# “Knowledge and Associated Factors Influencing Oral Care Provision Among ICU Nurses Caring for Mechanically Ventilated Patients at Tenwek Hospital, Kenya.”

**DOI:** 10.64898/2026.07.13.26357953

**Authors:** Shadrack Tuei, Vincent Kiprono Mukthar, Philip Towett

## Abstract

**Background:** Oral care is a critical nursing intervention for mechanically ventilated patients in intensive care units (ICUs) because it reduces oral microbial colonization and contributes to the prevention of ventilator-associated pneumonia (VAP), a major cause of morbidity, prolonged hospitalization, and increased healthcare costs among critically ill patients. Despite the availability of evidence-based guidelines, gaps in nurses’ knowledge and inconsistent implementation of oral care practices remain challenges, particularly in low- and middle-income settings.

**Objective:** This study aimed to assess the level of knowledge among ICU nurses regarding oral care for mechanically ventilated patients at Tenwek Hospital, Kenya, determine factors influencing knowledge, and examine the relationship between nurses’ knowledge and selected demographic and professional characteristics.

**Methods:** An analytical cross-sectional study design was employed. The study was conducted among ICU nurses at Tenwek Hospital. A sample of 38 nurses was selected from a target population of 60 ICU nurses using simple random sampling. Data were collected using a structured self-administered questionnaire assessing socio-demographic characteristics, knowledge of evidence-based oral care practices, awareness of guidelines, and factors influencing knowledge. Data were analyzed using descriptive statistics, chi-square tests, Pearson correlation, and logistic regression, with statistical significance set at p<0.05.

**Results:** Thirty-five nurses participated, yielding a response rate of 92.1%. The findings demonstrated generally high knowledge levels regarding oral care practices for mechanically ventilated patients. Most nurses correctly identified recommended oral care frequency (88.6%), oral assessment before care (91.4%), suctioning before and after oral care (94.3%), and the role of oral care in preventing VAP (97.1%). However, knowledge gaps were identified in areas such as toothbrushing practices and documentation of oral care procedures. Training, continuing professional education, availability of guidelines, workload, resources, supervision, and teamwork were identified as important factors influencing knowledge. The findings revealed that professional qualification (p=0.02), ICU experience (p=0.030) and formal oral care training (p=0.021) were significantly associated with nurses’ knowledge of oral care practices.

**Conclusion:** ICU nurses at Tenwek Hospital demonstrated adequate knowledge of evidence-based oral care practices for mechanically ventilated patients. However, targeted educational interventions are required to address identified gaps and strengthen standardized oral care delivery. Continuous professional development, consistent use of guidelines, supportive supervision, and adequate resource provision are recommended to enhance patient safety and reduce preventable ICU complications such as VAP.

## INTRODUCTION

Oral care is a fundamental component of nursing management for mechanically ventilated patients in the intensive care unit (ICU). Critically ill patients requiring mechanical ventilation are at increased risk of oral colonization and infection because endotracheal and tracheostomy tubes impair normal airway defense mechanisms, reduce mucociliary clearance, and promote accumulation of pathogenic microorganisms (Jun, 2022). Consequently, oral care serves not only to maintain patient comfort and hygiene but also as an important infection prevention intervention, particularly in reducing the risk of ventilator-associated pneumonia (VAP) (WHO, 2021).

Ventilator-associated pneumonia remains one of the most common healthcare-associated infections among mechanically ventilated patients and is associated with increased morbidity, mortality, prolonged ICU admission, extended mechanical ventilation, and increased healthcare costs. The accumulation of dental plaque and contaminated oral secretions creates a reservoir for microorganisms that may migrate into the lower respiratory tract through aspiration. Evidence indicates that consistent implementation of evidence-based oral care practices reduces microbial burden, lowers VAP risk, and improves outcomes among critically ill patients (Sarangi, Simon, & Sarangi, 2024).

ICU nurses are primarily responsible for providing continuous bedside care, including oral hygiene management for mechanically ventilated patients. Effective oral care requires adequate knowledge of evidence-based practices, including oral assessment, appropriate frequency of care, use of recommended oral care agents, suctioning techniques, and infection prevention measures. Nurses’ knowledge is essential in guiding clinical decisions, ensuring adherence to protocols, and promoting safe patient care (Masengesho, 2023).

Despite the availability of evidence-based guidelines, implementation of recommended oral care practices remains inconsistent worldwide. Studies indicate that a considerable proportion of mechanically ventilated patients develop VAP, with approximately one-quarter of ventilated patients affected in some settings (WHO, 2021). Even in healthcare systems with established protocols, adherence to oral care recommendations varies, while low- and middle-income countries experience additional challenges including limited training opportunities, inadequate resources, weak institutional support, and insufficient access to updated guidelines (Alqaissi & Qtait, 2025).

In sub-Saharan Africa, oral care practices among ICU nurses remain variable and are often not standardized. Although nurses recognize oral care as an important component of critical care, evidence suggests persistent knowledge gaps regarding evidence-based recommendations, resulting in inconsistent clinical practice (Narbutaitė et al., 2023). Studies from Rwanda, Nigeria, and South Africa have similarly reported inadequate knowledge of standardized oral care protocols despite nurses acknowledging its importance in preventing complications among mechanically ventilated patients (Masengesho, 2023; Kanamori et al., 2025).

In Kenya, oral care is recognized as an important infection prevention measure within ICU settings; however, limited empirical evidence exists regarding ICU nurses’ knowledge of oral care practices for mechanically ventilated patients. In many healthcare facilities, oral care delivery may depend on individual experience rather than standardized evidence-based approaches. Factors such as inadequate professional training, staffing challenges, competing clinical priorities, and limited access to updated protocols may contribute to variations in practice (WHO, 2021).

At Tenwek Hospital, preliminary observations suggest variability in ICU nurses’ understanding and application of evidence-based oral care practices for mechanically ventilated patients. Although oral care guidelines and standard operating procedures are available, their consistent utilization and understanding among nurses remain uncertain. Inadequate knowledge may contribute to inconsistent oral care delivery, increasing the risk of preventable complications such as VAP and other healthcare-associated infections.

Despite global recommendations and institutional protocols, there is limited documented evidence regarding ICU nurses’ knowledge of oral care practices for mechanically ventilated patients at Tenwek Hospital. This knowledge gap limits the ability to develop targeted educational and quality improvement interventions. Therefore, this study aimed to assess ICU nurses’ knowledge regarding oral care for mechanically ventilated patients at Tenwek Hospital, Kenya, identify factors influencing knowledge, and examine the relationship between nurses’ knowledge and selected demographic and professional characteristics.

The study was guided by the following specific objectives:

1. To determine the level of knowledge among ICU nurses regarding oral care practices for mechanically ventilated patients at Tenwek Hospital.
2. To assess factors influencing ICU nurses’ knowledge of oral care practices for mechanically ventilated patients at Tenwek Hospital.
3. To evaluate the relationship between ICU nurses’ knowledge and selected demographic and professional characteristics, including training, years of experience, and educational level.

## 2.0 LITERATURE REVIEW

### 2.1 Level of ICU Nurses’ Knowledge on Oral Care

Oral care is an essential nursing intervention for mechanically ventilated patients, aimed at maintaining oral hygiene, reducing microbial colonization, and preventing ventilator-associated pneumonia (VAP), a major ICU-acquired infection associated with increased morbidity and mortality (WHO, 2021; Tembo, 2016). Mechanically ventilated patients are particularly vulnerable to infection due to impaired airway defense mechanisms and accumulation of oral secretions, making nurses’ knowledge of evidence-based oral care practices critical for safe and effective care.

Knowledge of oral care involves understanding oral assessment, appropriate frequency of care, use of antiseptic agents such as chlorhexidine, suctioning techniques, infection prevention measures, and adherence to recommended protocols. Evidence indicates that nurses with adequate knowledge are more likely to recognize the importance of oral care and implement preventive measures effectively (Masengesho, 2023). However, studies globally reveal variations in ICU nurses’ knowledge, with many demonstrating general awareness but limited understanding of specific evidence-based recommendations (Kanamori et al., 2025).

In low- and middle-income countries (LMICs), knowledge gaps are more pronounced due to limited access to continuing professional development, inadequate training opportunities, and inconsistent institutional support (Alqaissi & Qtait, 2025). Studies from Africa, including Rwanda, Nigeria, and other settings, have reported that nurses often rely on routine practices or personal experience rather than standardized guidelines, contributing to inconsistent oral care delivery (Asadi & Jahanimoghadam, 2024; Masengesho, 2023). In Kenya, available evidence suggests that although ICU nurses understand the importance of oral care, gaps remain regarding current evidence-based practices and standardized protocols (Amutalla, 2023; Kumar et al., 2023).

### 2.2 Factors Influencing Nurses’ Knowledge of Oral Care

Several professional and institutional factors influence ICU nurses’ knowledge of oral care among mechanically ventilated patients. Educational preparation, previous oral care training, continuing professional development, and access to updated clinical guidelines are consistently identified as important determinants of knowledge (WHO, 2022; Narbutaitė et al., 2023).

Lack of structured training programs and limited opportunities for professional development contribute significantly to knowledge deficits. Studies in African healthcare settings have shown that inadequate institutional support and absence of standardized training negatively affect nurses’ ability to acquire and apply current oral care knowledge (Tefera et al., 2022; Asadi & Jahanimoghadam, 2024). In Kenya, additional challenges include limited access to training opportunities, inadequate oral care protocols, workload pressures, and resource constraints, which may reduce nurses’ ability to update and apply evidence-based knowledge consistently (Ochoki, 2022; Masengesho, 2023).

### 2.3 Relationship Between Nurses’ Knowledge and Practice Context

Evidence demonstrates a positive relationship between nurses’ knowledge and the quality of oral care provided to mechanically ventilated patients. Nurses with higher levels of knowledge are more likely to demonstrate improved adherence to evidence-based oral care procedures and infection prevention practices (Kanamori et al., 2025). However, knowledge alone may not guarantee optimal practice, as implementation is influenced by contextual factors such as availability of supplies, staffing levels, workload, leadership support, and organizational culture.

Studies from African settings indicate that nurses with better knowledge provide more appropriate oral care; however, institutional barriers may limit translation of knowledge into practice (Bwalya, 2025; Tefera et al., 2022). Similarly, Kenyan studies suggest that despite awareness of oral care importance, gaps in resources, training, and standardized systems contribute to inconsistent application of recommended practices (Amutalla, 2023). Therefore, improving oral care outcomes requires strengthening both nurses’ knowledge and the healthcare environment supporting practice.

### 2.4 Summary of Literature and Research Gap

The reviewed literature highlights the importance of ICU nurses’ knowledge in ensuring effective oral care and preventing complications such as VAP among mechanically ventilated patients. However, evidence demonstrates variability in knowledge levels across healthcare settings, with LMICs experiencing greater challenges due to limited training opportunities, inadequate resources, and weak institutional support.

In Kenya, particularly in rural and faith-based hospitals such as Tenwek Hospital, limited research has examined ICU nurses’ knowledge of oral care practices for mechanically ventilated patients. Existing studies have mainly focused on urban healthcare facilities, leaving insufficient evidence regarding knowledge levels and influencing factors in rural ICU settings.

Additionally, limited studies have integrated individual, professional, and institutional factors within a single framework to explain variations in nurses’ knowledge. This study therefore seeks to address this gap by assessing ICU nurses’ knowledge of oral care for mechanically ventilated patients and identifying factors influencing knowledge at Tenwek Hospital, Kenya.

The conceptual framework assumes that nurses’ knowledge directly influences their ability to provide evidence-based oral care, including oral assessment, infection prevention practices, suctioning, and use of appropriate oral care agents. However, this relationship is influenced by moderating factors such as education level, ICU experience, training exposure, availability of standard operating procedures, staffing, workload, and availability of supplies.

### 2.5 Conceptual Framework

The conceptual framework is guided by Benner’s Novice-to-Expert Theory, which explains the development of clinical competence through experience and professional growth, and Orem’s Self-Care Deficit Nursing Theory, which emphasizes the nurse’s role in meeting patients’ self-care needs when critically ill patients are unable to perform essential activities independently (Benner, 1984; Orem, 1971). Improved knowledge and supportive practice environments are expected to enhance oral care quality, reduce microbial colonization, and contribute to improved patient outcomes, including reduced VAP incidence and shorter ICU stays.

**Figure 1:**
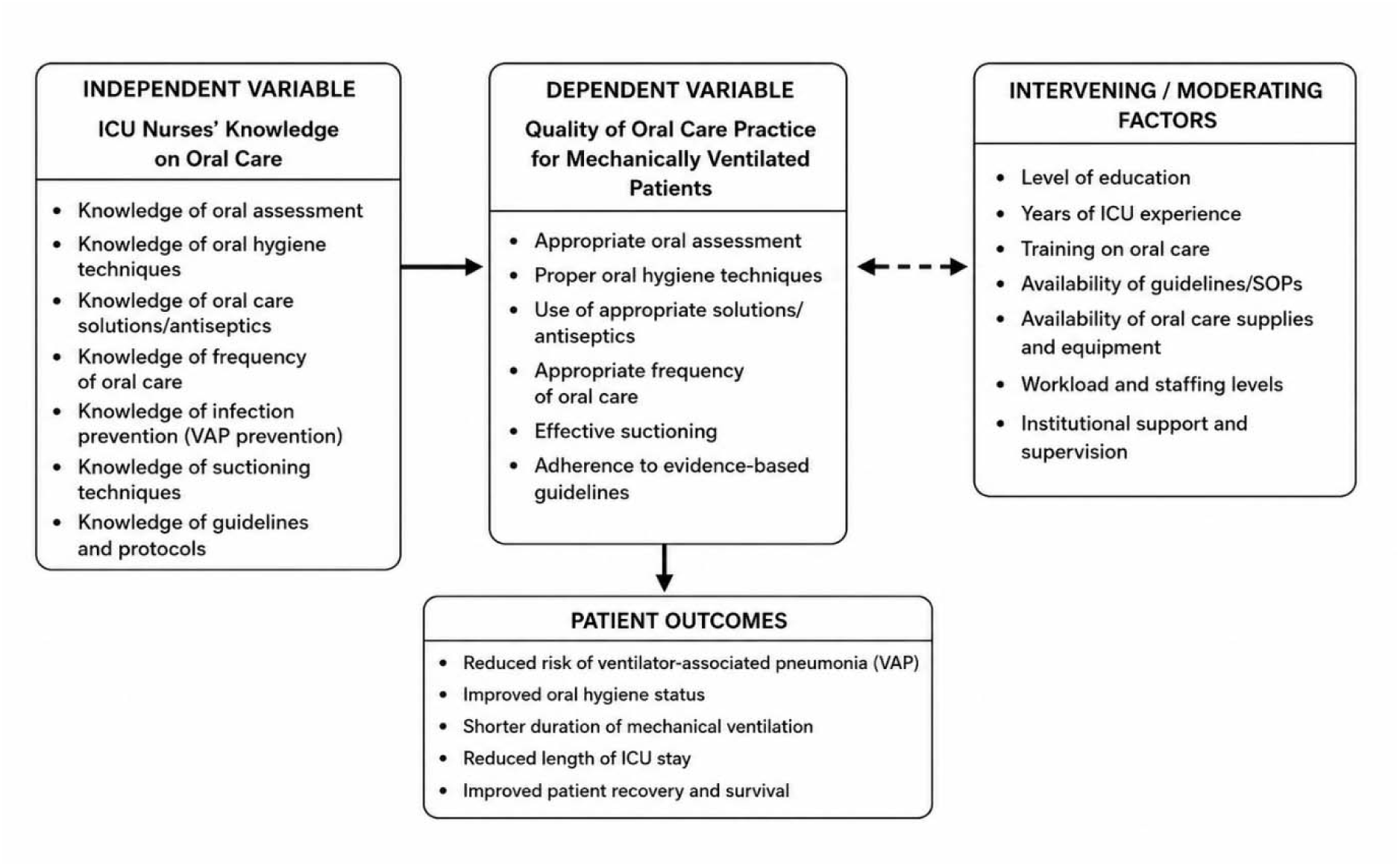
Conceptual Framework

## METHODS

### 3.1 Research Design

The study employed an analytical cross-sectional research design to assess ICU nurses’ knowledge of oral care for mechanically ventilated patients and determine factors associated with knowledge. The design was appropriate because it enabled data collection at a single point in time without manipulation of variables, providing a snapshot of nurses’ knowledge levels and associated factors within the ICU setting (Creswell & Creswell, 2018).

### 3.2 Study Setting

The study was conducted at Tenwek Hospital, a faith-based referral hospital located in Bomet County, Kenya. The hospital provides specialized healthcare services, including intensive care management for critically ill patients requiring mechanical ventilation. The ICU was selected because nurses provide continuous bedside care to mechanically ventilated patients and are responsible for implementing essential interventions such as oral care.

### 3.3 Study Population

The target population comprised registered nurses working in the ICU at Tenwek Hospital. According to hospital records (2025), approximately 60 ICU nurses were employed in the unit. The accessible population included ICU nurses available during the data collection period.

Participants were eligible if they were registered nurses working in the ICU, provided direct care to mechanically ventilated patients, had at least six months of ICU experience, and provided informed consent. Nurses on leave, those in administrative roles, those with less than six months ICU experience, and those who declined participation were excluded.

### 3.4 Sampling Procedure and Sample Size

The sample size was determined using Slovin’s formula due to the relatively small study population. From an estimated population of 60 ICU nurses, a sample of 38 nurses was selected. Simple random sampling was used to ensure that all eligible nurses had an equal opportunity to participate and to minimize selection bias.

### 3.5 Data Collection Instruments

Data were collected using a structured self-administered questionnaire developed from relevant literature and aligned with the study objectives. The questionnaire consisted of four sections covering socio-demographic and professional characteristics, knowledge of oral care practices for mechanically ventilated patients, awareness of evidence-based oral care guidelines, and factors influencing nurses’ knowledge, including training, experience, workload, and availability of guidelines.

### 3.6 Validity and Reliability of Instruments

Content validity was established through expert review by specialists in critical care nursing, infection prevention, and research methodology. Feedback from experts was used to improve clarity, relevance, and alignment of the instrument with the study objectives and conceptual framework.

The questionnaire was pre-tested among ICU nurses at Kapkatet Hospital using approximately 10% of the sample size to assess clarity, feasibility, and consistency. Reliability was evaluated using Cronbach’s alpha coefficient through SPSS version 30. A coefficient of ≥0.70 was considered acceptable, indicating adequate internal consistency of the knowledge assessment tool.

### 3.7 Data Collection Procedures

Following ethical approval from the relevant institutional review bodies, National Commission of Science, Technology and Innovation (NACOSTI), and permission from Tenwek Hospital administration, data collection was conducted among eligible ICU nurses. Participants received information about the study objectives and procedures before providing written informed consent.

The questionnaire was self-administered in a quiet environment and completed within approximately 20–30 minutes to minimize disruption of clinical duties. Completed questionnaires were collected, coded, and securely stored to maintain confidentiality.

### 3.8 Data Analysis

Data were reviewed for completeness, coded, and entered into SPSS version 30 for analysis. Descriptive statistics, including frequencies, percentages, means, and standard deviations, were used to summarize participants’ characteristics and levels of knowledge regarding oral care practices.

Inferential analysis was conducted to examine factors associated with nurses’ knowledge. Chi-square tests were used to assess associations between knowledge levels and categorical variables, while Pearson correlation was applied to determine relationships between knowledge and continuous variables. Binary logistic regression analysis was performed to identify predictors of adequate knowledge while controlling for potential influencing factors. Statistical significance was determined at a p-value of <0.05.

### 3.9 Ethical Considerations

Ethical approval was obtained from the relevant institutional ethics committee and NACOSTI, with additional permission granted by Tenwek Hospital administration. Participation was voluntary, and written informed consent was obtained from all participants.

Confidentiality and anonymity were maintained through the use of coded questionnaires and secure data storage. Participants were informed of their right to withdraw from the study at any stage without consequences. Data were used strictly for academic purposes and handled according to ethical principles outlined in the Declaration of Helsinki.

## RESULTS

### 4.1 Introduction

This chapter presents the findings of the study on ICU nurses’ knowledge regarding oral care practices for mechanically ventilated patients at Tenwek Hospital. The findings are presented according to the study objectives and include descriptive and inferential analyses.

### 4.2 Response Rate

A total of 38 questionnaires were distributed to ICU nurses, of which 35 were completed and returned, yielding a response rate of 92.1%. This response rate was considered adequate for analysis and interpretation of the study findings.

### 4.3 Demographic and Professional Characteristics of Respondents

The study involved 35 ICU nurses. More than half of the respondents (51.4%) were aged between 25 and 34 years, while females constituted the majority (65.7%). Nearly half of the participants held diploma qualifications (48.6%), followed by bachelor’s degree holders (31.4%). Most respondents had 2–5 years of nursing experience (40.0%) and 1–5 years of ICU experience (54.3%). Additionally, 68.6% reported having received formal training in oral care for mechanically ventilated patients.

### 4.4 Level of Knowledge Among ICU Nurses Regarding Oral Care Practices for Mechanically Ventilated Patients

Knowledge was assessed using ten evidence-based oral care statements. Overall, the findings demonstrated a high level of knowledge among ICU nurses regarding oral care for mechanically ventilated patients.

Most respondents correctly identified that oral care should be provided every 2–4 hours (88.6%), oral assessment should be conducted before oral care (91.4%), and suctioning should be performed before and after oral care (94.3%). All respondents (100%) correctly recognized the need to use personal protective equipment during oral care procedures, while 97.1% acknowledged that oral care helps prevent ventilator-associated pneumonia (VAP).

Similarly, 85.7% correctly identified sterile water or saline as appropriate solutions for oral rinsing, while 82.9% recognized the role of chlorhexidine in reducing VAP. Furthermore, 80.0% correctly indicated that lip moisturizers should be used to prevent dryness and cracking.

Despite the generally high knowledge levels, some knowledge gaps were identified. Only 74.3% correctly recognized that toothbrushing remains necessary for intubated patients, while 68.6% correctly disagreed with the statement that documentation of oral care is unnecessary in ICU settings. These findings suggest that although nurses demonstrated good overall knowledge, deficiencies existed regarding oral care documentation and mechanical oral cleaning practices.

### 4.5 Factors Influencing ICU Nurses’ Knowledge of Oral Care for Mechanically Ventilated Patients

The study examined factors perceived by nurses to influence their knowledge of oral care practices.

Training emerged as an important factor influencing knowledge. Nearly three-quarters of respondents (74.2%) agreed or strongly agreed that they had received adequate training on oral care for intubated patients. Similarly, 91.4% agreed or strongly agreed that continuous professional education improved their confidence and knowledge regarding oral care practices.

Availability of institutional guidelines was also identified as an important factor. Approximately 62.9% of respondents agreed or strongly agreed that clear oral care guidelines were available within the ICU, suggesting that access to protocols contributes to knowledge acquisition and standardization of practice.

Work-related factors were also reported to influence knowledge and practice. A large proportion of respondents agreed or strongly agreed that workload and nurse-patient ratios affected oral care provision (88.6%), while 82.8% reported that time constraints limited the frequency of oral care delivery.

Availability of oral care supplies was another influential factor, with 82.9% indicating that access to oral care resources affected practice. Likewise, 80.0% agreed that shortages of resources limited effective oral care provision.

Supervision and monitoring were perceived as important for adherence to oral care protocols, with 68.6% of respondents indicating that inadequate supervision negatively influenced compliance. Furthermore, 85.7% agreed that collaboration among ICU staff improved consistency in oral care practices.

The findings suggest that training, continuing education, availability of guidelines, workload, resource availability, supervision, and teamwork are key factors influencing nurses’ knowledge and implementation of oral care practices.

### 4.6 Relationship Between ICU Nurses’ Knowledge and Selected Demographic and Professional Factors

Chi-square analysis was conducted to determine the relationship between nurses’ knowledge and selected demographic and professional characteristics.

The findings revealed that professional qualification, ICU experience, and formal oral care training were significantly associated with nurses’ knowledge of oral care practices.

Formal training in oral care demonstrated the strongest association with knowledge (χ² = 12.84, p = .002). Nurses who had received formal oral care training were significantly more likely to demonstrate adequate knowledge compared to those who had not received training.

Professional qualification was also significantly associated with knowledge (χ² = 8.92, p = .030), indicating that nurses with higher educational qualifications were more likely to possess adequate knowledge regarding evidence-based oral care practices.

Similarly, years of ICU experience showed a significant association with knowledge levels (χ² = 9.76, p = .021), suggesting that nurses with longer ICU experience demonstrated greater knowledge of oral care practices for mechanically ventilated patients.

However, no statistically significant relationships were found between knowledge and age (p = .279), gender (p = .196), years of general nursing experience (p = .140), shift pattern (p = .315), or current role in the ICU (p = .233).

These findings indicate that formal training, educational attainment, and ICU-specific experience are important determinants of nurses’ knowledge regarding oral care for mechanically ventilated patients.

### 4.7 Chapter Summary

The study found that ICU nurses at Tenwek Hospital generally possessed a high level of knowledge regarding oral care practices for mechanically ventilated patients. Knowledge was particularly high regarding infection prevention, oral assessment, suctioning procedures, and the use of personal protective equipment. However, gaps were identified in documentation practices and toothbrushing recommendations.

Training, continuous education, availability of guidelines, workload, resource availability, supervision, and teamwork were identified as key factors influencing nurses’ knowledge. Furthermore, professional qualification, ICU experience, and formal oral care training were significantly associated with knowledge levels, while age, gender, shift pattern, and years of general nursing experience showed no significant associations.

## 5.0 DISCUSSION

### 5.1 ICU Nurses’ Knowledge Regarding Oral Care Practices for Mechanically Ventilated Patients

The first objective sought to determine the level of knowledge among ICU nurses regarding oral care practices for mechanically ventilated patients at Tenwek Hospital. The findings demonstrated that the majority of respondents possessed adequate knowledge of evidence-based oral care practices. High proportions of nurses correctly identified key recommendations, including the need for oral care every 2–4 hours (88.6%), the role of chlorhexidine in reducing ventilator-associated pneumonia (VAP) (82.9%), the importance of oral assessment prior to care (91.4%), and the need for suctioning before and after oral care procedures (94.3%). Furthermore, nearly all respondents recognized that oral care contributes to the prevention of VAP (97.1%) and that personal protective equipment should be used during oral care procedures (100%).

These findings suggest that ICU nurses at Tenwek Hospital have a strong understanding of the principles underpinning evidence-based oral care for mechanically ventilated patients. This level of knowledge is encouraging given the established relationship between oral hygiene, reduction of oral microbial colonization, and prevention of VAP among critically ill patients. The findings support recommendations from international critical care guidelines, which identify oral care as an essential component of VAP prevention bundles.

The findings are consistent with studies conducted in other critical care settings. For example, Asadi and Jahanimoghadam (2024) reported that ICU nurses demonstrated satisfactory knowledge of oral care interventions, particularly regarding infection prevention and oral assessment. Similarly, Alqaissi and Qtait (2025) found that nurses working in intensive care settings possessed adequate knowledge of oral hygiene practices and their role in preventing complications among mechanically ventilated patients. The relatively high knowledge levels observed in the current study may partly be attributed to the high proportion of respondents who had received formal oral care training (68.6%).

Despite the generally positive findings, important knowledge deficits were identified. Approximately one-quarter of respondents (25.7%) incorrectly believed that toothbrushing was unnecessary for intubated patients. This finding is clinically significant because toothbrushing remains one of the most effective methods of mechanical plaque removal and is recommended in many evidence-based oral care protocols. Similar misconceptions have been reported in previous studies conducted in low- and middle-income settings, where nurses often prioritize antiseptic mouth rinses while underestimating the importance of mechanical cleaning.

Another notable gap concerned documentation practices. Nearly one-third of respondents failed to recognize the importance of documenting oral care procedures. Documentation is a critical aspect of nursing practice because it facilitates continuity of care, accountability, communication among healthcare providers, and monitoring of patient outcomes. The persistence of this knowledge gap suggests that educational interventions should not only focus on procedural aspects of oral care but also emphasize the importance of documentation as part of comprehensive patient management.

Overall, the findings indicate that ICU nurses at Tenwek Hospital possess generally high levels of knowledge regarding oral care practices for mechanically ventilated patients. However, the identified gaps in toothbrushing and documentation suggest opportunities for targeted educational interventions aimed at strengthening adherence to evidence-based oral care standards.

### 5.2 Factors Influencing ICU Nurses’ Knowledge of Oral Care Practices

The second objective sought to assess factors influencing ICU nurses’ knowledge of oral care for mechanically ventilated patients. The findings demonstrated that both individual and organizational factors play important roles in shaping nurses’ knowledge levels.

Training emerged as one of the most influential factors affecting nurses’ knowledge. More than two-thirds of respondents reported receiving formal training in oral care, while the majority agreed that continuous professional education enhanced their confidence and understanding of oral care practices. These findings are consistent with previous studies indicating that structured educational programs significantly improve nurses’ knowledge of evidence-based oral care interventions. Educational interventions have been shown to increase awareness of current guidelines, improve adherence to recommended practices, and promote consistency in care delivery.

The findings support Benner’s Novice-to-Expert Theory, which proposes that professional competence develops through a combination of formal education and experiential learning. Nurses who receive continuous training are more likely to update their knowledge and integrate evidence-based recommendations into clinical practice. Consequently, investment in regular in-service training and continuing professional development programs remains essential for maintaining high standards of oral care in critical care settings.

The availability of clear oral care guidelines was also identified as an important factor influencing nurses’ knowledge. Nearly two-thirds of respondents agreed that the presence of hospital guidelines facilitated oral care provision. Clinical guidelines serve as important knowledge translation tools by converting research evidence into practical recommendations that can be readily applied in patient care. Previous studies have similarly reported that nurses working in units with standardized oral care protocols demonstrate greater knowledge and better adherence to evidence-based practices than those working in settings without formal guidelines.

Workload and time constraints were also perceived as important barriers. More than four-fifths of respondents reported that workload pressures affected the frequency and quality of oral care provision. Although workload primarily affects practice, excessive workload may also limit opportunities for learning, participation in training programs, and engagement with evidence-based updates. Similar findings have been reported in critical care settings where staffing shortages and competing clinical priorities reduce opportunities for knowledge acquisition and professional development.

Resource availability emerged as another significant factor. The majority of respondents indicated that access to oral care supplies influenced their ability to provide appropriate care. Resource limitations may hinder the practical application of acquired knowledge and create discrepancies between what nurses know and what they are able to implement. Previous studies have identified shortages of oral care supplies, chlorhexidine solutions, and suction equipment as barriers to evidence-based oral care implementation.

The findings further revealed that supportive supervision and multidisciplinary collaboration contribute positively to nurses’ knowledge development. Nurses who receive regular supervision, feedback, and mentorship are more likely to remain informed about current standards of care and maintain compliance with recommended practices. Likewise, collaborative practice environments encourage knowledge sharing and professional learning among healthcare team members.

Collectively, these findings suggest that nurses’ knowledge is influenced not only by individual educational preparation but also by organizational factors such as training opportunities, availability of guidelines, resource adequacy, supportive supervision, and collaborative workplace cultures.

### 5.3 Relationship Between ICU Nurses’ Knowledge and Selected Demographic and Professional Factors

The third objective sought to evaluate the relationship between ICU nurses’ knowledge and selected demographic and professional factors, including training, years of experience, and educational level. The findings demonstrated that knowledge was significantly associated with several professional characteristics, particularly formal training, educational qualification, and ICU experience.

Formal training showed the strongest association with knowledge-related outcomes. Nurses who had received oral care training were significantly more likely to demonstrate adequate knowledge compared to those without training. This finding aligns with previous studies indicating that structured educational programs improve nurses’ understanding of evidence-based oral care principles and promote adherence to clinical guidelines. Training provides opportunities for updating knowledge, correcting misconceptions, and reinforcing best practices.

Professional qualification was also significantly associated with knowledge. Nurses with higher educational attainment demonstrated better knowledge levels than those with lower qualifications. This finding is consistent with the literature suggesting that advanced nursing education enhances critical thinking, evidence appraisal skills, and the ability to apply research findings in clinical practice. Nurses with bachelor’s and postgraduate qualifications may have greater exposure to evidence-based practice concepts during their professional preparation, thereby strengthening their knowledge of specialized interventions such as oral care for mechanically ventilated patients.

Years of ICU experience were similarly associated with higher knowledge levels. Nurses with longer ICU experience demonstrated better understanding of oral care practices than less experienced colleagues. This finding supports Benner’s theoretical proposition that expertise develops through repeated clinical exposure and experiential learning. Over time, ICU nurses encounter diverse patient care situations that reinforce theoretical knowledge and facilitate the development of clinical judgment.

In contrast, no statistically significant relationships were observed between knowledge and age, gender, shift pattern, or current nursing role. These findings suggest that professional exposure and educational opportunities may be more important determinants of knowledge than personal demographic characteristics. Similar observations have been reported in previous critical care studies, where knowledge levels were more strongly influenced by training and professional development than by demographic variables.

Further evidence supporting the importance of knowledge was demonstrated by the significant relationship between knowledge and oral care performance. Nurses with adequate knowledge consistently reported better implementation of oral assessment, oral suctioning, patient positioning, and infection prevention measures than those with inadequate knowledge. These findings suggest that knowledge serves as a foundational prerequisite for evidence-based practice and reinforces the importance of educational interventions in improving quality of care.

In summary, the findings indicate that formal training, higher educational attainment, and greater ICU experience are key determinants of nurses’ knowledge regarding oral care for mechanically ventilated patients. These results underscore the need for healthcare institutions to strengthen educational programs, promote continuing professional development, and support mentorship initiatives aimed at enhancing knowledge and improving patient outcomes in critical care settings.

## 6.0 CONCLUSION AND RECOMMENDATIONS

### 6.1 CONCLUSION

The study found that ICU nurses at Tenwek Hospital had generally adequate knowledge of evidence-based oral care practices for mechanically ventilated patients. Most respondents correctly identified recommended oral care procedures, including oral assessment, suctioning, infection prevention measures, and the role of oral care in preventing ventilator-associated pneumonia. However, gaps were noted in knowledge related to toothbrushing and documentation, indicating the need for targeted educational reinforcement.

Nurses’ knowledge was influenced by both individual and organizational factors. Formal training, continuous professional education, availability of oral care guidelines, supportive supervision, teamwork, and adequate resources facilitated knowledge acquisition and application. Conversely, workload pressures, staffing constraints, resource shortages, and limited supervision hindered effective knowledge utilization.

The study established that nurses’ knowledge was significantly associated with professional qualification, ICU experience, and formal oral care training. Nurses with higher educational attainment, greater ICU experience, and formal training demonstrated better knowledge levels. In contrast, age, gender, shift pattern, and current role in the ICU were not significantly associated with knowledge. These findings underscore the importance of specialized training and critical care experience in strengthening nurses’ knowledge of oral care practices.

### 6.2 Recommendations

The study came up with the following recommendations;

1. Strengthen Continuous Education and Training Tenwek Hospital should provide regular in-service training and continuing professional development on evidence-based oral care, focusing on identified knowledge gaps such as toothbrushing and documentation.
2. Standardize Oral Care Guidelines The hospital should ensure the availability and consistent use of evidence-based oral care protocols within the ICU to promote standardized practice and knowledge retention.
3. Enhance Institutional Support Management should strengthen supervision, mentorship, teamwork, and ensure adequate oral care resources while addressing staffing and workload challenges that may hinder adherence to recommended practices.
4. Support Professional Development Structured orientation and mentorship programs should be provided for newly recruited and less experienced ICU nurses to strengthen knowledge and promote evidence-based oral care practices.

## Data Availability

All data produced in the present study are available upon reasonable request to the authors

## REFERENCES

1. Al-Jubouri, M. B. A., & Jaafar, S. A. (2021). Nurses’ Knowledge and Practice Toward Oral Care for Intubated Patients. Indian Journal of Public Health Research & Development, 9(9).

2. Alqaissi, N., & Qtait, M. (2025). Knowledge, Attitudes, and Practices of Intensive Care Unit Nurses Regarding Oral Care for Intubated Patients in Hebron Hospitals, Palestine. SAGE Open Nursing, 11, 23779608251368041.

3. Amutalla, S. (2023). Nurses Practices on Closed Endotracheal Tube Airway Suctioning at KNH Adult Critical Care Unit (Doctoral dissertation, University of Nairobi).

4. Asadi, N., & Jahanimoghadam, F. (2024). Oral care of intubated patients, challenging task of ICU nurses: a survey of knowledge, attitudes and practices. BMC Oral Health, 24(1), 925.

5. Benner, P. (1984). From novice to expert: Excellence and power in clinical nursing practice. Addison-Wesley.

6. Bwalya, R. (2025). Compliance of nurses with nursing care protocols for ventilated patients in the University Teaching Hospital’s adult intensive care unit in Lusaka, Zambia (Doctoral dissertation, The University of Zambia).

7. Creswell, J. W., & Creswell, J. D. (2018). Research design: (5th ed.). Sage Publications

8. Kanamori, D., Fujii, T., Yoshida, M., Ito, R., Sakai, A., Takahashi, H., & Tochio, T. (2025). Oral quality of care for intubated patients in the intensive care unit: examination o f bacterial count and microbiota. Critical Care, 29(1), 320.

9. Kumar, S., Singh, B., Mahuli, A. V., Kumar, S., Singh, A., & Jha, A. K. (2023). Assessment of Nursing Staff’s Knowledge, Attitude and Practice Regarding Oral Hygiene Care in Intensive Care Unit Patients: A Multicenter Cross-sectional Study. Indian Journal of Critical Care Medicine: Peer-reviewed, Official Publication of Indian Society of Critical Care Medicine, 28(1), 48.

10. Masengesho, F. (2023). Nurses’ knowledge and practice of oral care for intubated patients in selected referral and teaching hospitals in Rwanda (Doctoral dissertation, University of Rwanda).

11. Narbutaitė, J., Skirbutytė, G., & Virtanen, J. I. (2023). Oral care in intensive care units: Lithuanian nurses’ attitudes and practices. Acta Odontologica Scandinavica, 81(5), 408–413.

12. Orem, D. E. (1971). Nursing: Concepts of practice. McGraw-Hill.

13. Tembo, E. (2016). Intensive care nurses’ knowledge, attitudes and practices of oral care for patients with oral endotracheal intubation (Master’s thesis, University of the Witwatersrand, Johannesburg (South Africa)).

14. Wang, Z., Zhou, Y., & Guo, J. (2020). Oral health and prevention of ventilator-associated pneumonia in critically ill patients. BMC Oral Health, 20, 354. 10.1186/s12903-020-01335-9

15. World Health Organization. (2021). Essential emergency and critical care: a consensus among global clinical experts.

16. World Health Organization. (2022). Oral health. Retrieved from https://www.who.int/news-room/fact-sheets/detail/oral-health

